# Targeted detection of microbes in synbiotic medical foods SBD111 and SBD121 to evaluate gut persistence: a randomised, open label trial

**DOI:** 10.1101/2025.08.07.25333216

**Authors:** Kelsey J. Miller, Ian M. Wolff, Luis Arturo Montes de Oca Valeriano, Maria J. Soto-Giron, Sushrut Jangi, Eric M. Schott, Mark R. Charbonneau, Alicia E. Ballok, Gerardo V. Toledo

## Abstract

The viability and persistence of orally administered microbes in the human gut are essential to their biological function. We previously described the development of two synbiotic medical foods, SBD111 and SBD121, each comprising four food-derived microbial strains and prebiotic fibers for the dietary management of postmenopausal bone loss and rheumatoid arthritis, respectively. Here, we report a randomised, open-label clinical study examining the persistence of SBD111 and SBD121 microbes in faecal samples from healthy adults following administration for seven days. Thirty-eight participants, aged 18-64 years with BMI 18.5-35 kg/m^2^, were randomised to receive one of the two synbiotic medical foods daily for one week, followed by a four-week monitoring period. Employing quantitative PCR (qPCR), shotgun metagenomics, and culture-based assays, we evaluated the presence and viability of the microbial strains comprising each synbiotic medical food during and after administration. SBD111 and SBD121 were well-tolerated with minimal adverse events reported. Strains were detected in over 80% of participants during the administration period, with strain abundance peaking in the first week. Persistence in the follow-up period varied by strain and detection method. The microbial strains were detected by qPCR and metagenomic sequencing for a median of seven days and three days during the follow-up period, respectively. However, *Bacillus amyloliquefaciens* was consistently detected for seven days by both methods. Culture-based assays confirmed the presence of viable strains from both synbiotic medical foods in stool samples up to one-week post-consumption. Faecal metagenome diversity and metabolic functional potential remained stable throughout the administration and follow-up periods, aligning with previous findings in healthy adults. Collectively, these results establish that SBD111 and SBD121 deliver viable microbes that transiently persist in the gut, reinforcing their promise for safe and targeted dietary interventions.

This trial is registered at ClinicalTrials.gov (NCT06614166) and was funded by Solarea Bio Inc.

## 1. Introduction

Probiotics are characterised as living organisms that, when administered in sufficient quantity, can be beneficial to the host (Hill et al., 2014). The most well-known probiotic microbes are bacteria, such as *Bifidobacteria* and *Lactobacillus* species, but yeasts, including *Saccharomyces* species, have also been shown to confer health benefits (Staniszewski and Kordowska-Wiater, 2021). Several studies have reported increased probiotic survival in simulated gastric environments with coadministration of prebiotics, non-digestible food components that promote microbial viability and activity (Peredo et al., 2016; Sathyabama et al., 2014; Pandey et al., 2015). The combination of probiotics and prebiotics are collectively referred to as synbiotics.

A critical factor for orally administered microbes to confer health benefits is the ability to persist in the gastrointestinal (GI) tract (Millette et al 2008), a trait that varies substantially by strain (Masco et al., 2007; Elo et al., 1991). Persistence is commonly assessed using molecular and culture-based detection methods that enable strain-specific tracking in complex microbial communities, such as stool samples (Morelli and Pellegrino, 2021). Once administered, these strains must compete with resident microbes for limited nutrients and ecological niches, which can influence their gut persistence and metabolic function (Pandey et al., 2015). Evidence suggests that strain persistence is typically transient, lasting days to weeks after intake stops (Tremblay et al., 2023; Maldonado-Gómez et al., 2016). Microbial colonisation dynamics are influenced by several host-specific factors, including microbiome composition, diet, and immune factors that contribute to high interindividual variability (Zmora et al., 2018). Given the transient nature of microbial colonisation and these host-specific factors, evaluating microbial persistence has become essential to understanding the function of orally administered microbial interventions.

We previously described the development of two rationally designed synbiotic medical foods, SBD111 and SBD121, formulated for the dietary management of postmenopausal bone loss and rheumatoid arthritis (RA), respectively (Lawenius et al., 2022; Easson et al., 2022; Easson et al., 2024). Both formulations contain probiotic strains isolated from a variety of healthy foods sources, combined with prebiotic fibers, oligofructose and blueberry powder, and encased in enteric capsules to support microbial viability and activity through the GI tract. In a recently completed randomised clinical study of 286 early postmenopausal women, twice daily administration of SBD111, containing *Leuconostoc mesenteroides, Lactiplantibacillus plantarum, Pichia kudriavzevii*, and *Levilactobacillus brevis* reduced bone loss in participants with osteopenia, elevated body mass index (BMI), and those with elevated body fat (Schott et al., 2025). SBD121, containing *Lactococcus lactis, Schleiferilactobacillus harbinensis, Bacillus amyloliquefaciens*, and *Levilactobacillus brevis*, is currently under investigation in a randomised, placebo-controlled efficacy trial of recently diagnosed RA patients in combination with the standard of care treatment, methotrexate (ClinicalTrials.gov ID NCT06005220). A previous study to evaluate safety and tolerability of SBD111 in healthy participants demonstrated no detection of the orally administered microbial strains after a 28-day washout period (Sahni et al., 2023). However, the kinetics of microbial abundance and persistence in the faecal microbiome, as well as the viability of these strains *in vivo*, following administration of these synbiotic medical foods remained to be determined.

Here, we report the results of an open-label study designed to assess the abundance and persistence of the microbial constituents of medical foods SBD111 and SBD121 in stool samples of 36 healthy volunteers over time, using a combination of culture-based methods, quantitative polymerase chain reaction (qPCR), and metagenomic sequencing. Secondary objectives included characterization of changes in gut microbial diversity and metabolic pathways, and the exploratory endpoint assessed viability of the synbiotic microbes. By integrating multiple detection methods, this study aims to accurately elucidate SBD111 and SBD121 microbial persistence, viability, and effects on the endogenous microbiome in the human gut.

## 2. Materials and methods

### Study design and participants

An open-label study was conducted at a single site in Waltham, Massachusetts, United States, between June and December 2024, to investigate the detection and persistence of medical food strains in faecal samples during and following oral administration of SBD111 and SBD121 in healthy adults. Secondary objectives included characterization of changes in gut microbial diversity and functional potential over time. Exploratory analyses assessed viability of probiotic strains in stool by recovering live isolates at baseline, Day 7, and Day 14. This trial was approved by the Institutional Review Board at Advarra (food trial to evaluate the kinetics of the medical foods SBD111 and SBD121 persistence in healthy adults, IRB# Pro00078728, ClinicalTrials.gov ID NCT06614166) and enrolled 38 healthy adults aged 18-64 years agreeing to follow study procedures (Figure 1).

**Figure 1.**
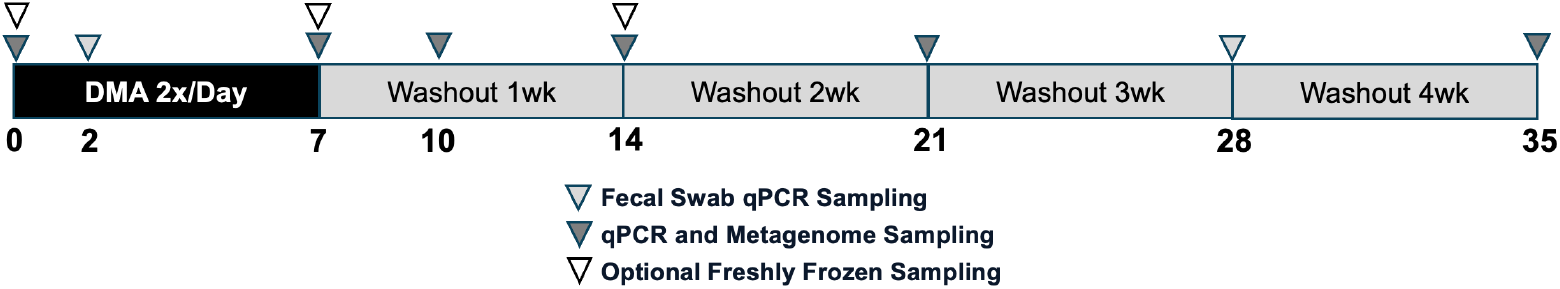
Study design schematic. Participants were randomised to receive either SBD111 or SBD121 for a seven-day intervention period, followed by a four-week wash-out period. Triangles indicate the faecal sample collection timepoints, annotated by the type of analysis performed (e.g., live microbial enumeration, qPCR, and metagenomics).

Participants were recruited through online advertisements and flyers. Interested individuals completed an online screening survey, followed by a virtual eligibility assessment conducted by study staff. Participants were eligible if they had with a body mass index (BMI) between 18.5 and 35 kg/m^2^, were in general good health as determined by screening within 30 days prior to study start (see supplemental methods for exclusion criteria) and were willing to comply with the study protocol and availability requirements throughout the study duration. Participants were not asked to change their diet other than to refrain from taking probiotic supplements. Consumption of probiotic-containing foods such as fermented dairy products and vegetables was permitted. All participants provided written informed consent before the study was conducted. Each participant was randomised 1:1 into one of two study groups to receive either SBD111 or SBD121 using centralized, computer-generated randomization. Due to the open-label design, neither participants nor investigators were blinded to group assignments. A clinical monitor provided oversight of study conduct and participant safety throughout the trial.

### Study procedure and intervention

The intervention consisted of four daily capsules. SBD111 capsules contained *Levilactobacillus brevis* (7.5 × 10^9^ CFU), *Lactiplantibacillus plantarum* (7.5 × 10^9^ CFU), *Leuconostoc mesenteroides* (7.5 × 10^9^ CFU), and *Pichia kudriavzevii* (1.25 × 10^9^ CFU), while SBD121 capsules contained *Levilactobacillus brevis* (7.5 × 10^9^ CFU), *Schleiferilactobacillus harbinensis* (3.5 × 10^9^ CFU), *Lactococcus lactis* (7.5 × 10^9^ CFU), and *Bacillus amyloliquefaciens* (7.5 × 10^9^ CFU). Capsules also included oligofructose and dried blueberry powder, in addition to inert flow aids, magnesium stearate and silicon dioxide (Table S1).

Study participants collected faecal DNA swabs (4N6 FLOQSwabs; Copan Diagnostics, Murrieta, CA, USA) at baseline (Day 0) and on Days 2, 7, 10, 14, 21, 28, and 35 to assess microbial prevalence and persistence. Swabs were stored at room temperature prior to DNA extraction. To evaluate viability of medical food strains before and after administration, a subset of participants (12 receiving SBD111 and 17 receiving SBD121) provided additional faecal samples (∼1 g) on Days 0, 7, and 14, which were immediately frozen by participants and transported to the study site on dry ice for culture-based analysis. Adherence to the supplementation protocol was monitored by providing participants with an excess number of capsules and assessing capsule counts during the Day 7 follow-up interview, where participants reported the number of remaining capsules.

### Medical food strain detection by quantitative polymerase chain reaction (qPCR)

Total DNA was extracted from each faecal swab sample using the ZymoBIOMICS DNA miniprep kit (Zymo Research, Irvine, CA, USA) according to manufacturer’s instructions. Purified PCR products of each strain using the strain-specific primers were combined per group at 1^10^ copies per strain, including the 16S purified product and specificity was checked by gel electrophoresis. Standard curves were generated by 10-fold serial dilutions of the mixture. Each faecal DNA sample was run in duplicate and normalised to the lowest concentration (above 2 ng/µL) among the eight samples collected from each participant. If DNA concentrations were less than or equal to 2 ng/µL, samples were run in triplicate undiluted.

Absolute quantification of strains in faecal samples was performed by qPCR on a CFX96 Touch Real-Time thermocycler (Bio-Rad Laboratories, Inc, Hercules, CA, USA) using the primers and probes listed in Table S2. Each reaction was performed with 10 µL volumes using the PrimeTime Gene Expression Master Mix (Integrated DNA Technologies, Inc., Coralville, IA, USA), strain primers (0.25 µM), strain probes (0.25 µM) and DNA template (1 µL). The PCR programme consisted of 3 min at 95°C, followed by 40 cycles of 95°C for 10 sec and 63°C for 30 sec.

### Shotgun metagenomics and microbiome analysis

Stool samples for metagenomic analysis were collected from study participants on Days 0, 7, 10, 14, 21, and 35. Metagenomic DNA libraries were prepared using the Illumina DNA Prep (M) Tagmentation Kit (Illumina Inc., San Diego, CA, USA), along with IDT Illumina DNA/RNA UD Index Sets A and B, following the manufacturer’s protocol. Library concentrations were quantified using a Qubit 3 Fluorometer (Thermo Fisher Scientific, Waltham, MA, USA). Libraries were then pooled at equimolar concentrations, and the final pooled library was quantified using both Qubit 3 Fluorometer and real-time PCR. Fragment size distribution was assessed using an Agilent 2100 Bioanalyser to ensure quality and consistency. Sequencing was conducted by Novogene Co., Ltd. (Beijing, China) on an Illumina NovaSeq X Plus instrument, utilizing a 25B Flow Cell and paired-end 150 bp (PE150) chemistry. Each sample was sequenced to a target depth of 5 Gb of data. Sequencing of samples from Day 14 was performed in a separate batch, which introduced variability in read number. Due to these batch effects, Day 14 samples were excluded from microbial community profiling analyses.

Raw sequencing reads were quality trimmed and residual adaptors were removed using Trimmomatic v0.39 (Bolger et al., 2014) with the following parameters: -phred33 LEADING:3 TRAILING:3. To eliminate host contamination, reads were aligned to the human reference genome (GRCh37, GCF_000001405.13) using Bowtie2 v2.5.4 (Langmead and Salzberg, 2012), and alignments were processed with SAMtools v1.13 (Li et al., 2009) to filter human-origin sequences. All metagenomic samples were subsequently subsampled to a depth of 10 million read pairs. Three samples with fewer than 10 million read pairs were retained as they exhibited acceptable microbial community coverage (greater than 75% as estimated using Nonpareil v3.5.5; Rodriguez-R et al., 2018).

Taxonomic profiling of metagenomic samples was conducted using MetaPhlAn4 (Blanco-Míguez et al., 2023), employing the mpa_vJun23_CHOCOPhlAnSGB_202307 reference database. Taxa detected in at least 5% of samples were retained for downstream analysis. Functional profiling of microbial communities was performed using HUMAnN3 (Beghini et al., 2021) with default settings. Functional annotation utilised multiple reference pathway databases, including UniRef, KEGG, UniPathway, and MetaCyc. The resulting pathway abundance output was filtered to retain pathways with an equal to or greater than 5% prevalence across samples.

### SBD111 and SBD121 detection in stool metagenomes

To monitor the presence of SBD111 and SBD121 strains in participant stool metagenomes across study timepoints, custom strain-specific k-mers were developed using Strainer2 (Aggarwala et al., 2021) and further refined using in-house bioinformatics pipelines. Target genomes were fragmented into overlapping 31 bp k-mers then screened against a custom reference database of public and internal human stool metagenomic datasets, genomes of closely related strains with an Average Nucleotide Identity (ANI) greater than 98%, and the MetaPhlAn4 marker gene database (mpa_vJun23_CHOCOPhlAnSGB_202307) (Blanco-Míguez et al., 2023).

K-mer frequencies were calculated across this reference database to identify informative, strain-specific k-mers. Informative k-mers were defined as the least prevalent kmers in the database, comprising 1% of total k-mers for *L. brevis, L. plantarum, L. mesenteroides, L. lactis*, and *S. harbinensis*, 2% of total k-mers for *B. amyloliquefaciens*, and 23% of total k-mers for *P. kudriavzevii*. For each sample, the presence of a strain was determined by mapping the set of informative k-mers to the metagenomic reads and calculating k-mer coverage, defined as the proportion of unique strain-specific k-mers detected in the sample relative to the total number of k-mers in the reference set. A strain was considered present if the k-mer coverage exceeded 0.03× for bacterial strains or 0.01× for fungal strains.

### Viable microbial detection using selective plating and qPCR identification

Samples were transferred to an anaerobic chamber (Coy Laboratories, Grass Lake, MI, USA), weighed, and processed into 10% (w/v) faecal suspensions in phosphate-buffered saline (PBS). Suspensions were homogenized, filtered through 100 µm cell strainers, serially diluted, and plated on both selective and non-selective media (Table S3) and incubated for 48 hrs before enumerating colony forming units (CFUs). Following incubation, 24 colonies (Day 0) or 32 colonies (Days 7 and 14) for each sample were picked from these media (except brucella), lysed in Tris-EDTA buffer (boiled at 95°C for 30 min, centrifuged at 5000 rpm for 1 min), and used for qPCR-based strain identification. Modified fluorophores were used for *L. lactis* and *L. plantarum* probes (Table S2). *P. kudriavzevii* and *B. amyloliquefaciens* were detected using iTaq Universal SYBR Green Supermix (Bio-Rad, Hercules, CA, USA).

### Statistical analysis

Metagenomic sample statistical analyses were conducted in R Statistical Software (v4.3.2; R Core Team 2021). Relative abundance values were square root-transformed. Changes in repeated measures variables across study groups were assessed using linear mixed-effects models (LMER). The models included random effects to account for repeated measurements within subjects and fixed effects for repeated measures, e.g., time points. Differential abundance of taxa and pathways was evaluated using LMER as implemented in the lmerTest package (v3.1.3; Kuznetsova et al., 2017).

To control multiple hypothesis testing, p-values were adjusted using the Benjamini-Hochberg false discovery rate (FDR) correction. Adjusted p-values < 0.05 were considered statistically significant. Changes in richness and Shannon diversity over time for both taxa and pathways were also assessed with the linear mixed-effects model implementation from lmerTest. P-values less than 0.05 were considered statistically significant.

qPCR data for strain abundance in stool samples were first analysed for outliers using the Grubbs test, followed by a mixed-effects model and Tukey’s multiple comparisons test in GraphPad Prism version 10.1.0 (Waltham, MA, USA). Time to non-detection via qPCR was assessed using a one-way ANOVA, while the plating results from selective agar were analysed with a two-way ANOVA; both were followed by Tukey’s multiple comparisons tests.

## 3. Results

### SBD111 and SBD121 are well tolerated in healthy adults

Thirty-eight individuals were recruited and randomised to receive either SBD111 or SBD121. One participant did not initiate the study, and another withdrew after one week; data from these individuals were excluded from subsequent analyses. The remaining 36 participants (18 per group) completed the study. The ages and body mass index (BMI) of study participants of each group were similar (Table 1). Capsule consumption adherence was high (98.3% average). However, one participant randomised to the SBD121 group consumed the probiotic prior to providing their baseline (Day 0) sample.

**Table 1.** Participant demographics and reported adverse events.

| Medical food | SBD111 | SBD121 |
| --- | --- | --- |
| **Participation** |  |  |
| Recruited | 20 | 18 |
| Initiated (%) | 19 (95%) | 18 (100%) |
| Completed (%) | 18 (90%) | 18 (100%) |
| **Clinical Characteristics** |  |  |
| Age (+/- SD) | 31.9 (9.1) | 33.4 (10.8) |
| BMI (+/- SD) | 25.3 (4.0) | 24.7 (4.5) |
| **Sex/Gender** |  |  |
| Male (%) | 10 (50%) | 10 (55.6%) |
| Female (%) | 10 (50%) | 7 (38.9%) |
| Other | 0 (0%) | 1 (5.6%) |
| **Race/Ethnicity** |  |  |
| White | 14 (70%) | 13 (72.2%) |
| Asian | 1 (5.0%) | 1 (5.6%) |
| Mixed | 1 (5.0%) | 4 (22.2%) |
| Other/Unknown | 4 (20%) | 0 (0%) |
| Hispanic/Latino | 7 (35%) | 8 (44.4%) |
| **Adverse Events (AEs)** |  |  |
| Total AEs | 7 | 3 |
| Number of participants with AEs | 6 | 2 |
| Mild Severity | 4 | 3 |
| Moderate Severity | 3 | 0 |
| Gas and/or Bloating | 3 | 2 |
| Diarrhoea | 1 | 0 |
| Headache | 1 | 0 |
| Upper Respiratory Infection | 2 | 1 |

The incidence of adverse events was minimal, with seven reported for SBD111 (four mild, three moderate) and three for SBD121 (three mild) (Table 1). No severe adverse events were reported. The most commonly reported adverse events were mild gastrointestinal symptoms such as gas and bloating. A total of 10 adverse events were reported across eight participants. However, no participants discontinued participation in the study due to adverse events. Some events, including upper respiratory infections, were assessed as unlikely to be related to the intervention.

### Microbe detection and persistence in human faeces via quantitative polymerase chain reaction (qPCR)

qPCR was employed using DNA extracted from faecal swabs collected throughout the study to evaluate the presence and persistence of the medical food strains following oral administration. Custom-designed primers and probes enabled sensitive detection of each target microbe over time.

Analysis of baseline stool samples from the SBD111 group revealed detectable levels of *L. plantarum* and *L. brevis* in one and six participants, respectively. Similarly, *L. lactis, B. amyloliquefaciens*, and *L. brevis* were detected in baseline stool samples from the SBD121 group for one, two, one, and two participants, respectively. Following administration, all strains from both synbiotic medical foods were detected in over 80% of participants. Notably, by Day 7, *L. plantarum, L. lactis, B. amyloliquefaciens*, and *L. brevis* from both synbiotics were detected in 100% of participants who received their respective formulations (Table 2). *L. mesenteroides* and *S. harbinensis* were detected in 94% of their respective participant groups and *P. kudriavzevii* was detected in 83% of SBD111-consuming participants. *P. kudriavzevii, L. lactis, B. amyloliquefaciens*, and the SBD121 *L. brevis* each showed a significant increase in abundance by Day 2 compared to Day 0, while *L. plantarum* and *L. mesenteroides* exhibited a significant increase in abundance from Day 0 to Day 7 (Figures 2A and B). Significant increases in abundance of *S. harbinensis* in the SBD121 group and *L. brevis* in the SBD111 group were not observed. Each of the lactic acid bacterial strains and *B. amyloliquefaciens* demonstrated a significant decrease in abundance by Day 14, seven days after consumption of the medical food was discontinued.

**Table 2.** Strain detection frequencies per detection method on day seven.

| Microbe | qPCR | Metagenomics | Microbial plating |
| --- | --- | --- | --- |
| **SBD111 group** |  |  |  |
| *L. mesenteroides* (SBS04255) | 94% | 78% | 75% |
| *L. plantarum* (SBS04260) | 100% | 72% | 67% |
| *P. kudriavzevii* (SBS04263) | 83% | 50% | 50% |
| *L. brevis* (SBS04254) | 100% | 67% | 100% |
| **SBD121 group** |  |  |  |
| *L. lactis* (SBS04916) | 100% | 100% | 35% |
| *S. harbinensis* (SBS04913) | 94% | 50% | 0% |
| *B. amyloliquefaciens* (SBS04877) | 100% | 100% | 100% |
| *L. brevis* (SBS04254) | 100% | 94% | 100% |

**Figure 2.**
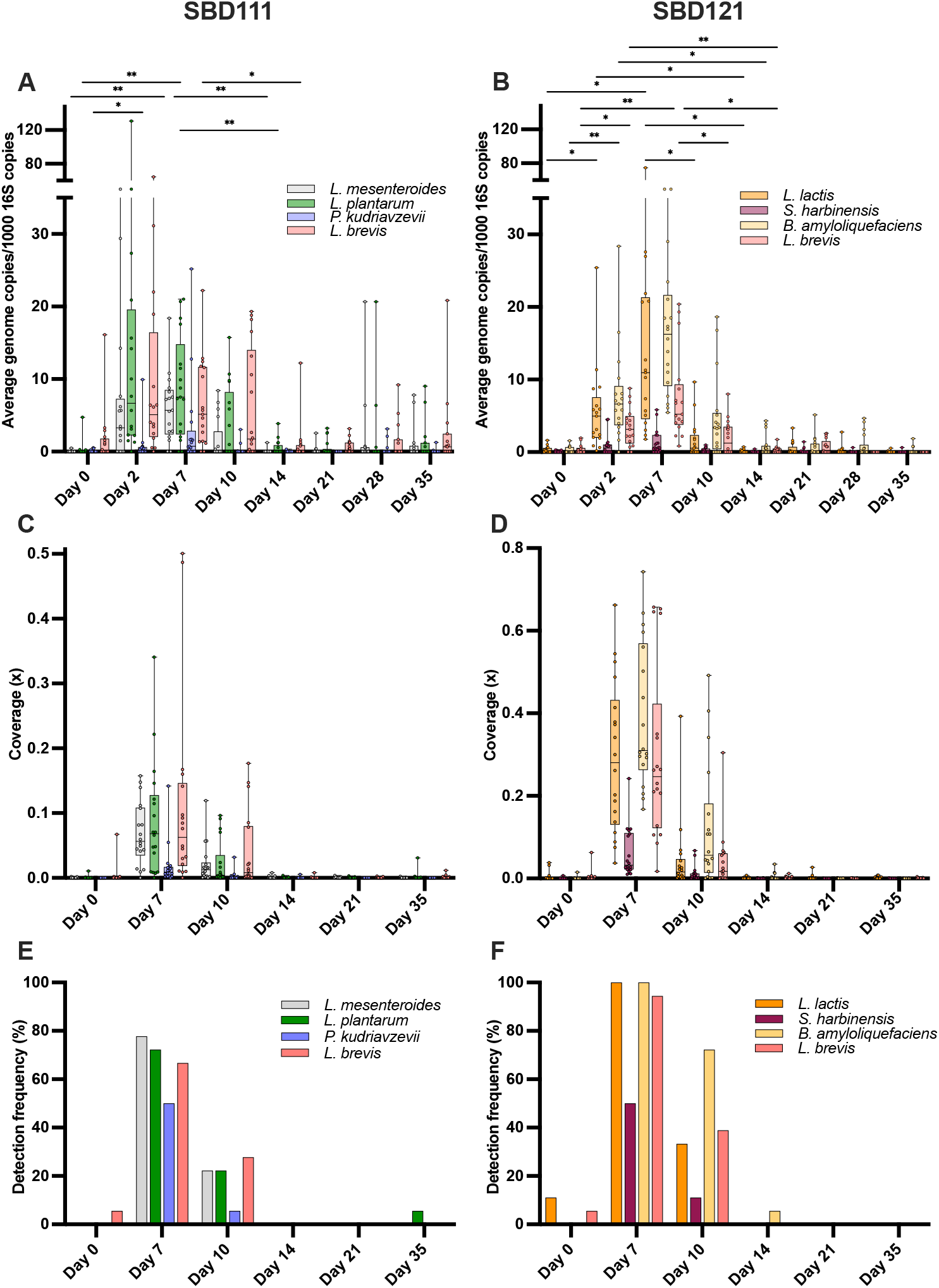
Detection and persistence of probiotic strains in stool samples. **A-B.** Quantification of SBD111 and SBD121 microbes in faecal samples by qPCR. Data are presented as mean values with distribution shown using box plots. Statistical analyses were performed using a mixed-effects model with Tukey’s multiple comparison test. Significant differences between time points are indicated (*p < 0.05 and **p < 0.01). **C-D**. Coverage of genome-specific k-mer sets for each microbial strain comprising SBD111 (*L. plantarum, L. brevis, L. mesenteroides*, and *P. kudriavzevii*) and SBD121 (*L. brevis, B. amyloliquefaciens, S. harbinensis*, and *L. lactis*) in stool metagenomes of participants at baseline, 7 days, 14 days, 10 days, 21 days, and 35 days. Box plots represent the median, interquartile range (IQR), and 95% confidence intervals. Each point corresponds to an individual stool metagenome sample. **E-F**. Detection frequency of SBD111 and SBD121 strains based on the genome-specific k-mer sets identified in stool metagenomes of participants for each time point.

To further characterize strain persistence, the duration of detectability following product discontinuation was examined. The median time to non-detection was seven days post-administration for strains *L. plantarum*, and *L. brevis* in the SBD111 group and *L. lactis* in the SBD121 group, while *B. amyloliquefaciens* and the SBD121 *L. brevis* exhibited persistence beyond one week (11 and 14 days, respectively). Additionally, *S. harbinensis* exhibited a median time to non-detection of six days. The time to non-detection did not differ significantly between strain pairs, except for *P. kudriavzevii* and *L. brevis* in the SBD111 group, which were detected for a median of three- and seven-days post ingestion, respectively (Figure S1).

### Medical food strain detection and persistence in human faeces via shotgun metagenomic sequencing

As a complement to qPCR-based detection, shotgun metagenomic sequencing was used to detect the administered medical food strains and determine their persistence in the gastrointestinal tract of healthy adults. To achieve this, genome-specific k-mer sequences were used to identify each microbial strain in stool metagenomes. Analysis of baseline stool samples confirmed the absence of medical food strains in all participants, except for one individual randomised to the SBD121 group, who showed detectable levels of *L. lactis* and *L. brevis*, likely attributable to product consumption prior to sample collection, as reported by the participant. Additionally, *L. lactis* was detected in a second participant from the SBD121 group at baseline, and *L. brevis* was identified in one participant from the SBD111 group.

Genome coverage analysis revealed that all administered strains were detected in at least one participant at Day 7 in both groups, with SBD121 strains exhibiting higher k-mer coverage than SBD111 strains (Figure 2C and D). Strain coverage peaked at Day 7, declined by Day 10, and was undetectable at later time points. Notably, two exceptions were observed: *L. plantarum* was detected in one participant on Day 35, and another participant had *B. amyloliquefaciens* detected on Day 14.

The detection frequency of the medical food microbes on Day 7 varied across groups (Figure 2E and F). In the SBD111 group, *L. mesenteroides* and *L. plantarum* were detected in over 70% of participants, *L. brevis* in 66%, and *P. kudriavzevii* in 50%. In contrast, in the SBD121 group, *B. amyloliquefaciens* and *L. lactis* were detected in all participants, *L. brevis* in 94%, and *S. harbinensis* in 50%.

### Analysis of microbial diversity and metabolic function during and after consumption of synbiotic medical foods

To explore whether the presence of medical food strains was associated with changes in faecal microbial diversity, shifts in microbial species and metabolic pathway abundance in stool metagenomes during and after product consumption were assessed. No significant differences in species Shannon diversity were observed between Day 7 and any other time point in either group. However, species richness significantly increased in the SBD111 group from Day 7 to Day 10 (p<0.05 LMER) (Figure S2A and B). No significant changes were detected in the Shannon diversity index or richness of metabolic pathways across time points in either group (Figure S3A and B).

To further assess changes at the species and functional pathway levels, stool metagenomes were analysed using MetaPhlAn for taxonomic profiling and HUMAnN for functional pathway analysis. Consistent with the strain-level analysis described earlier, a non-significant increase was observed in the relative abundance of *L. plantarum* and *L. brevis* in the SBD111 group on Day 7 compared to other time points (Figure S4A). Notably, *L. mesenteroides* showed a statistically significant increase on Day 7 relative to Day 21. In contrast, in the SBD121 group, all strains showed a significant increase at Day 7 compared to other time points, with the exception of *S. harbinensis* (Figure S4B). LMER analysis also identified four metabolic pathways with significantly increased abundance in the SBD121 group (Table S4). On Day 7, PWY-6992 (1,5-anhydrofructose degradation) and PWY-3001 (superpathway of L-isoleucine biosynthesis I) were significantly elevated compared to Day 21. Additionally, at Day 7 versus Day 35, CENTFERM-PWY (pyruvate fermentation to butanoate) and PWY-6590 (superpathway of *Clostridium acetobutylicum* acidogenic fermentation) were significantly increased. No significant pathway-level differences were observed in the SBD111 group.

### Viable medical food strain detection in human faeces via selective plating

To demonstrate the viability of the medical food strains after consumption and transit through the gastrointestinal tract, additional one-gram stool samples provided by participants who volunteered on days 0, 7, and 14 were homogenized and plated onto selective and non-selective media and analysed using colony qPCR. No significant changes in total microbial abundance were observed across faecal samples in either group or media type (Figure S5). In both groups, MRS agar with antibiotics displayed a small, non-significant increase in total colony counts by Day 7. Similarly, M17 agar and TSA supplemented with cycloheximide exhibited non-significant increases in microbial abundance, corresponding to increases in *L. lactis* and *B. amyloliquefaciens* respectively, in the SBD121 group.

To complement abundance measurements and further evaluate strain viability, detection frequency and positive identification rates were analysed via colony qPCR to capture how consistently each microbial strain was recovered across participants and time points. By Day 7, all strains comprising SBD111 and SBD121 were detected in at least one faecal sample, with the exception of *S. harbinensis* (Figure 3A). Positive identification rates ranged from 0.67% to 100% across strains and days (Figure 3B). Notably, *L. brevis* was detected in 100% of participants on Day 7 in both groups, with high positive identification rates. On Day 0, prior to supplementation, *L. brevis* was detected in a low percentage of participants in both groups. By Day 14, although detection remained variable, *L. brevis* was more frequently identified compared to baseline. *B. amyloliquefaciens* was detected in all participants on Day 7, with viability detected in over 50% of individuals through Day 14. Both *L. mesenteroides* and *L. plantarum* were present at all time points but exhibited consistently low positive identification rates. *L. lactis* was recovered from 35% of participants on Day 7, with a low identification rate and persistence observed in only one participant by Day 14. *P. kudriavzevii* was detected exclusively on Day 7 in 50% of participants, with a 100% identification rate in those samples.

**Figure 3.**
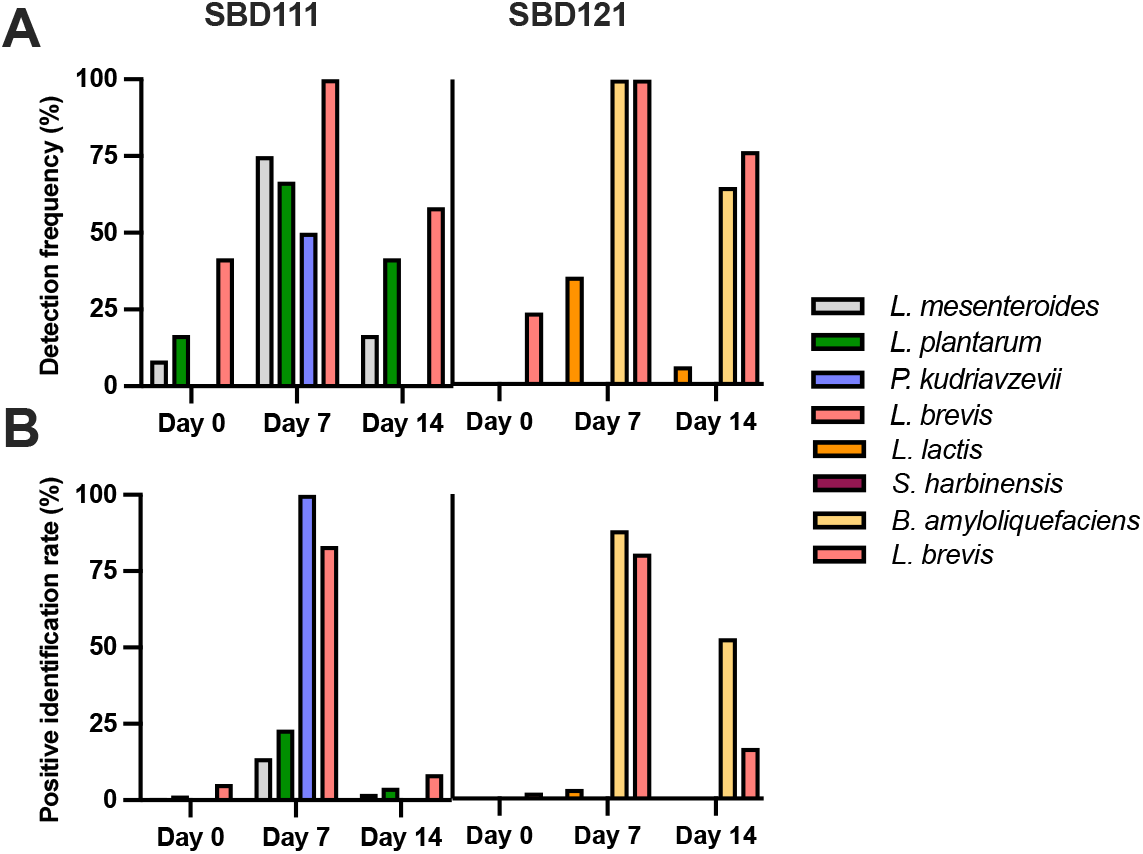
Culture-based frequency and positive identification rate of administered probiotic strains. **A.** Detection frequency reflects the proportion of participants in which a viable representative microbe was recovered from stool. **B**. Positive identification rate reflects the proportion of cultured confirmed to match the intended strain. Bars indicate the proportion of participants in which each probiotic strain was detected by plating methods and the frequency was confirmed by quantitative PCR across study timepoints. Data are presented as percentages (n = 12 and 17 participants per SBD111 and SBD121 groups, respectively).

## 4. Discussion and conclusion

This open-label study evaluated the presence, viability, and persistence of strains in the medical foods SBD111 and SBD121 following daily consumption by healthy adults. Using culture-based and molecular methods, we demonstrated that these strains remained detectable and viable in stool for a limited period post-administration. These findings provide direct evidence that certain microbial strains can transiently persist in the gut, a property believed to be important for exerting functional effects. The use of enteric-coated capsules likely enhanced survival by protecting strains from gastric acid and promoting targeted release in the lower small intestine (Kim et al., 2016). While prior studies have reported persistence ranging from five to 30 days (Morelli and Pellegrino, 2021; Hanifi et al., 2015; Maldonado-Gomez et al., 2016), few have employed a targeted, multi-platform detection strategy to validate strain-specific presence. Our combined use of qPCR, metagenomics, and microbiological methods offers a robust framework for evaluating strain-specific viability and persistence following ingestion.

Probiotic persistence in the gut varies widely by strain, detection method, delivery format, and host (Morelli and Pellegrino, 2021). A previous study reported a median time to non-detection of *L. rhamnosus* GG (LGG) of 17 days, and another study having detectable LGG in several participants at the study end (Saxelin et al., 2010). In this study, qPCR detected several strains, including *L. plantarum* and *L. lactis*, up to seven days post-supplementation in some participants, while *L. brevis* and *B. amyloliquefaciens* persisted beyond one week in the SBD121 group. Notably, *L. brevis* persisted one week longer in SBD121 participants than in SBD111 individuals, suggesting possible synergistic interactions within the SBD121 formulation that enhance survival of *L. brevis*. Metagenomic sequencing supported overall patterns of persistence but revealed shorter detection durations, three days post consumption, except for *B. amyloliquefaciens*, which remained detectable at Day 7. Notably, culture-based methods diverged from molecular results: live *B. amyloliquefaciens* was recovered from over half of participants at Day 14, suggesting that viable cells may persist longer than indicated by DNA-based methods alone. These discrepancies reflect differences in sensitivity and specificity between detection platforms and highlights the complex, host-dependent nature of probiotic persistence.

Interestingly, several administered strains were detected at baseline in multiple participants, a phenomenon reported in prior studies and likely attributable to dietary sources (i.e. cheese, yogurt, fermented foods) or the presence of genetically similar resident microbes that cannot be distinguished by current methods (Dommels et al., 2009; Hanifi et al., 2015; Saxelin et al., 2010). In our study, *L. brevis, L. plantarum*, and *L. lactis* were identified at baseline by both qPCR and metagenomics, possibly due to the unrestricted consumption of lactic acid bacteria-rich foods (Zapasnik et al., 2022). Detection of *S. harbinensis* and *P. kudriavzevii* was limited by sparse reference genomes, while high genomic similarity complicated resolution of *B. amyloliquefaciens* from related strains. *L. plantarum* and *L. brevis* are also known commensals (Matsuda et al., 2009), raising the possibility of their native presence. However, significant increases in detection frequency post-intervention support the interpretation that these signals reflect the administered strains rather than endogenous populations. These findings emphasise the ongoing challenge of differentiating administered from endogenous or dietary microbes, even with strain-specific approaches.

Live *S. harbinensis* was not recovered post-intervention using culture-based techniques, likely due to both methodological and biological limitations. The lower relative abundance of *S. harbinensis*, detected by qPCR and metagenomics, may be due to its lower initial concentration in the medical food and could suggest limited replication in the gut. Recovery of this organism in culture was further hindered by the strain’s limited natural antibiotic resistance and the absence of a highly selective medium. Additionally, competition from resident gut microbiota may have suppressed *S. harbinensis* growth, particularly given its slower replication rate. Similar challenges have been noted previously, with reduced detection in faecal compared to colonic samples (Alander et al., 1999), highlighting the importance of multi-platform approaches for evaluating probiotic persistence.

In parallel with evaluating strain-level persistence and viability, we assessed the broader impact of synbiotic administration on gut microbial community structure. Microbiome composition remained stable throughout the intervention, with no significant changes in Shannon diversity or species richness, indicating that a seven-day intake of SBD111 or SBD121 did not alter the resident microbial community. These findings align with the 28-day SBD111 safety study (Sahni et al., 2023) and prior studies showing no microbiome shifts following short-term probiotic supplementation in healthy individuals (Larson et al., 2006). Similarly, Mai et al. (2016) reported subject-specific microbiome clustering, highlighting inter-individual variation as a stronger determinant of microbiome composition than probiotic intake. Viable counts of total and lactic acid bacteria (LAB) also remained stable throughout the intervention, indicating no substantial change in cultivatable gut bacterial populations. These findings suggest that short-term synbiotic consumption does not significantly affect overall microbial diversity, species abundance, or functional pathways in the gut.

The observed differences in detection and viability between strains emphasise the necessity of strain-level tracking when evaluating probiotic formulations. The combined use of qPCR, metagenomic sequencing, and viability plating provided complementary insights: qPCR offered sensitive strain-specific detection, metagenomic sequencing confirmed strain-level presence within the community, and culture-based methods demonstrated viability. Concordance across these platforms strengthens the validity of our findings by addressing the limitations of individual methods, such as false positives from dietary or environmental sources, and minimizing misidentification of closely related taxa through strain-resolved sequencing (Morelli and Pellegrino, 2021). This study demonstrates that strains in SBD111 and SBD121 can be detected, remain viable, and persist transiently in the human gut for up to one-week post-consumption. These results highlight the need for continued administration to sustain high probiotic levels and inform future clinical and mechanistic studies.

## Acknowledgments

We would like to thank all study participants for their willingness to participate in the study. Additionally, thanks to Tony Tempesta and David Easson for providing study product materials.

## Author Contributions

Conceptualisation and clinical trial coordination, AEB; data acquisition and analysis, KJM, IMW, LAMdOV, MJSG, SJ, AEB; writing-original draft, KJM, IMW, MJSG, AEB; writing, reviewing, and editing, KJM, IMW, LAMdOV, MJSG, SJ, EMS, MRC, AEB, GVT. All authors read and approved the final manuscript.

## Conflict of interest

KJM, IMW, LAMdOV, MJSG, EMS, MRC, AEB, GVT are employed by Solarea Bio, Inc. SJ served as clinical monitor of the clinical study and was paid a consulting fee.

## Data availability

The data presented in this study are available on request from the corresponding author.

## Notes

### Clinical Trial

NCT06614166

### Funding Statement

This study was funded by Solarea Bio Inc.

### Author Declarations

Ethics committee/IRB of Advarra, Inc. gave ethical approval for this work

