## Supplementary Methods and Data for "Targeted detection of microbes in synbiotic medical foods SBD111 and SBD121 to evaluate gut persistence: a randomised, open label trial"

### Supplemental methods

**Table S1.** Composition of SBD111 and SBD121 medical foods

| Ingredient | Amount in each capsule (CFUs) | Daily amount in four capsules (CFUs) | Ingredient in each capsule (mg) |
|---|---|---|---|
| **SBD111 probiotic medical food formulation** | | | |
| *Levilactobacillus brevis* (SBS04254) | $7.5x10^{9}$ | $3.0\ x10^{10}$ | 31 |
| *Lactiplantibacillus plantarum* (SBS04260) | $7.5x10^{9}$ | $3.0\ x10^{10}$ | 38 |
| *Leuconostoc mesenteroides* (SBS04255) | $7.5x10^{9}$ | $3.0\ x10^{10}$ | 23 |
| *Pichia kudriavzevii* (SBS04263) | $1.25x10^{9}$ | $5.0x10^{9}$ | 92 |
| Oligofructose | NA | NA | 143 |
| Dried, ground blueberry powder | NA | NA | 143 |
| Magnesium stearate | NA | NA | 5 |
| Silicon dioxide | NA | NA | 5 |
| Total | $2.375x10^{10}$ | $9.5x10^{10}$ | 480 |
| **SBD121 probiotic medical food formulation** | | | |
| *Levilactobacillus brevis* (SBS04254) | $7.5\ x10^{9}$ | $3.0\ x10^{10}$ | 30 |
| *Schleiferilactobacillus harbinensis* (SBS04913) | $3.5\ x10^{9}$ | $1.4\ x10^{10}$ | 16 |
| *Lactococcus lactis* (SBS04916) | $7.5\ x10^{9}$ | $3.0\ x10^{10}$ | 17 |
| *Bacillus amyloliquefaciens* (SBS04877) | $7.5\ x10^{9}$ | $3.0\ x10^{10}$ | 11 |
| Oligofructose | NA | NA | 168 |
| Dried, ground blueberry powder | NA | NA | 168 |
| Magnesium stearate | NA | NA | 5 |
| Silicon dioxide | NA | NA | 5 |
| Total | $2.6x10^{10}$ | $1.04x10^{11}$ | 420 |
| Abbreviations: NA = not applicable | | | |

### Exclusion criteria

Participants were excluded if they had used probiotic or prebiotic supplements within 30 days prior to enrollment or were unwilling to avoid such supplements during the study; had known allergies to probiotics, maltodextrin, or berries; had recent antibiotic use (within two months); had undergone major intestinal surgery or endoscopy within three months; or had a history of drug or alcohol abuse. Additional exclusions included clinically significant systemic abnormalities, presence of indwelling catheters or feeding tubes, febrile illness or recent diarrhea within 72 hours before baseline, active gastrointestinal diseases or history of dysmotility, chronic liver disease or hepatitis, structural heart disease or prior endocarditis, immunosuppression (including HIV or transplant recipients), celiac disease, history of cancer (except non-melanoma skin cancer or cancers >10 years prior), autoimmune diseases treated with immunosuppressants, active tuberculosis, pregnancy or breastfeeding, cognitive impairment interfering with consent or adherence, bowel movement frequency less than one per 36 hours, recent participation in experimental trials within 60 days, or any other condition deemed by the investigator to jeopardize participant safety or study completion.

### Design of strain primers and probes

Primers for the SBD111 and SBD121 strains (Table S2) were designed based on genome-specific regions identified using the Fur (Finding Unique genomic Regions) tool (Haubold et al., 2021). A set of closely related genomes sharing an Average Nucleotide Identity (ANI) greater than 98% with the target strain was compiled and input into Fur to identify these regions. The tool was then used to detect genomic regions unique to each target strain by excluding sequences conserved across the related genomes. The resulting strain-specific sequences served as templates for primer design using Primer-BLAST (https://www.ncbi.nlm.nih.gov/tools/primer-blast/). The primer design parameters were set as follows: amplicon length of 200–250 base pairs (bp), primer length between 20–25 bp, and GC content ranging from 40–60%. For each strain, four primer pairs targeting distinct genomic regions were designed.

Within the unique genomic region amplified by the strain primers, TaqMan probes were designed for each probiotic microbe. Probes were designed using Benchling Molecular Biology tools (San Francisco, CA, USA) with sizes ranging from 19–33 bp. Each probe was synthesized with a 5' reporter dye (HEX, Cy5, Cy5.5, 6-FAM or TEX 615), a 3' quencher (Iowa Black FQ or RQ-Sp, Black Hole 3), and an internal quencher (ZEN or TAO). Specificity was confirmed in silico using BLAST against NCBI's non-redundant database. All primers and probes were purchased from Integrated DNA Technologies, Inc. (Coralville, IA, USA).

**Table S2.** Primers and probes used in this study

| Microbial strain | Primer sequence (5'-3') | Amplicon size (bp) | Probe sequence (5'-3') | Reference |
|---|---|---|---|---|
| *L. mesenteroides* (SBS04255) | GTTATTGCATCGGGTCAAAGAGC<br>CCTACCTGTGTTACGAACCATTTC | 232 | Cy5/ACTTTGTAG/TAO/CCTTCTTTAACGCGGTTATTATTG/3IAbRQSp | This study |
| *L. plantarum* (SBS04260) | TGGAAGCGTCGATGTTAGGT<br>TGCTTACTGTCGTACCTGCC | 201 | 5Cy55/AGGGTAGAAATAAGCCTAACAAGCGGC/3BHQ-3<br>5TEX615/AGGGTAGAAATAAGCCTAACAAGCGGC/3IAbRQSp | This study |
| *P. kudriavzevii* (SBS04263) | ACCGCTGTTAAATCCTCCTCTATG<br>GAATAGAATACGATTCTGTGCGGG | 203 | 56-FAM/AAAGGGAAA/ZEN/GCAGCGAAGAAAGA/3IABkFQ | This study |
| *L. lactis* (SBS04916) | GATGCCGATTCGAAAAGTGAAG<br>AGCATCTTCAAGACGTATTCCG | 234 | 5Cy55/ACTTGAATCTCTAATATGGGGCCTGGATTG/3IAbRQSp<br>5TEX615/ACTTGAATCTCATAATATGGGGCCTGGATTG/3IAbRQSp | This study |
| *S. harbinensis* (SBS04913) | TCAACTCCTTCGGGGAATGC<br>CGCTGGGTCTCTAGAAGCTG | 240 | 56-FAM/TCGCCCTCT/ZEN/TGAGCA TCGTTTGTCG/3IABkFQ | This study |
| *B. amyloliquefaciens* (SBS04877) | CCGACAGTTCTAATGCCGGA<br>GGCTCCACTCGGAAAAACGA | 249 | 5Cy5/AGCACCGAT/TAO/TAATGAAATGCCTGCCGC/3IAbRQSp | This study |
| *L. brevis* (SBS04254) | TAGTCATGGGGGTTAGACGTTATG<br>GAACACCTTGATACCCTCTTGTGAC | 246 | 5HEX/GCCAGGGAA/ZEN/TTAAGGTCATGCGGTATAAC/3IABkFQ | This study |
| All bacteria (16S) | CGGTGAATACGTTCYCGG<br>AAGGAGGTGATCCRGCCGCA | 174 | 5TEX615/CTTGTACACACCGCCCGTC/3IAbRQSp | BACT1369F, PROK1541R, and TM1389F (Suzuki et al., 2000) |

**Microbial whole-genome sequencing**

High-quality genomes were generated for SBD111 and SBD121 strains using a hybrid assembly approach with Illumina and Oxford Nanopore sequencing data. For Illumina sequencing, genomic DNA was extracted from pure cultures using the Zymo Quick-DNA Bacterial/Fungal Kit (Zymo Research, Irvine, CA, USA). DNA libraries were constructed with the Nextera XT DNA Library Preparation Kit (Illumina Inc., San Diego, CA, USA) following the manufacturer's instructions. Library concentration was measured using a Qubit 3.0 Fluorometer (Thermo Fisher Scientific, Waltham, MA, USA). Sequencing was performed on an Illumina MiSeq platform (MiSeq Control Software v2.6) with 2 × 250 bp paired-end reads. Raw reads were trimmed and filtered based on a Phred score >20 and a minimum fragment length of 50 bp using SolexaQA v3.1.7.1 (Cox et al., 2010).

For Oxford Nanopore sequencing, genomic DNA from *Pichia kudriavzevii* was extracted using the MasterPure Yeast DNA Purification Kit (Lucigen), and genomic DNA from bacterial strains was extracted using the Quick-DNA Fungal/Bacterial kit (Zymo Research, Irvine, CA, USA). DNA libraries were prepared with the Nanopore Genomic DNA by Ligation Kit, and sequencing was performed on the MinION platform (Oxford Nanopore Technologies, Oxford, UK) using MinKNOW software v20.10.3. Fast5 reads were basecalled and converted to Fastq format using Guppy v6.5.7 (dna_r10.4.1_e8.2_400bps_hac.cfg). Read quality was evaluated with NanoFilt v2.8.0 (De Coster et al., 2018), using a Phred score >20 and a minimum fragment length of 1,000 bp.

Filtered Nanopore reads were assembled using Flye v2.8.2 (Kolmogorov et al., 2019), and the assembly was polished with Medaka v1.8.1 (Oxford Nanopore Technologies, Oxford, UK). Illumina reads were mapped to the assembled contigs for three rounds of polishing using Pilon v1.23 (Walker et al., 2014) and BWA-MEM v0.7.17 (Li, 2013). Assembly quality was evaluated using QUAST v5.2.0 (Gurevich et al., 2013), and genome completeness using CheckM v1.1.3 (Parks et al., 2015) for bacterial genomes and BUSCO v5.5.0 (saccharomycetes_odb10 dataset) (Seppey et al., 2019) for the *P. kudriavzevii* genome.

**Table S3.** Media types for the isolation of synbiotic medical food strains

| Media | Strain target | Incubation conditions |
|---|---|---|
| Brucella | Total anaerobes | Anaerobic, 37°C |
| De Man–Rogosa–Sharpe (MRS) | Total lactic acid bacteria (LAB) | Anaerobic, 37°C |
| Potato dextrose agar (PDA) with 40 µg/mL chlortetracycline | *P. kudriavzevii* | Aerobic, 30°C |
| tryptic soy agar (TSA) with 50 µg/mL cycloheximide | *B. amyloliquefaciens* | Aerobic, room temperature |
| MRS with vancomycin and cefoxitin (4 µg/mL each) | *L. mesenteroides, L. plantarum, L. lactis, S. harbinensis,* and *L. brevis* | Anaerobic, 37°C |
| MRS with vancomycin and trimethoprim (4 µg/mL each) | *L. mesenteroides* | Aerobic, room temperature |
| M17 with 2 µg/mL chloramphenicol | *L. lactis* | Aerobic, room temperature |

**Table S4.** Metabolic pathways showing differential abundance in stool metagenomes of participants in the SBD121 group, comparing Day 7 to other time points (baseline, Day 10, 21, and 35).

**Day 7 vs. Day 21**

| Pathway | Abundance in SBD121 participants at Day 7 | | Abundance in SBD121 participants at Day 21 | | p-value (linear mixed effects regression) | p-value (linear mixed effects regression with FDR correction) |
|---|---|---|---|---|---|---|
| | Mean | SD | Mean | SD | | |
| PWY-6992: 1,5-anhydrofructose degradation | 0.0231 | 0.0211 | 0.0109 | 0.0206 | 2.44E-06 | 0.0011 |
| PWY-3001: superpathway of L-isoleucine biosynthesis I | 0.1709 | 0.0100 | 0.1639 | 0.0104 | 0.0001 | 0.0232 |

**Day 7 vs. Day 35**

| Pathway | Abundance in SBD121 participants at Day 7 | Abundance in SBD121 participants at Day 35 |
|---|---|---|

| | Mean | SD | Mean | SD | p-value (linear mixed effects regression) | p-value (linear mixed effects regression with FDR correction) |
|---|---|---|---|---|---|---|
| CENTFERM-PWY: pyruvate fermentation to butanoate | 0.0519 | 0.0107 | 0.0433 | 0.0118 | 0.0001 | 0.0361 |
| PWY-6590: superpathway of Clostridium acetobutylicum acidogenic fermentation | 0.0586 | 0.0117 | 0.0490 | 0.0130 | 0.0002 | 0.0361 |


### Supplemental figure legends

**Figure S1.** Time to non-detection of probiotic strains.

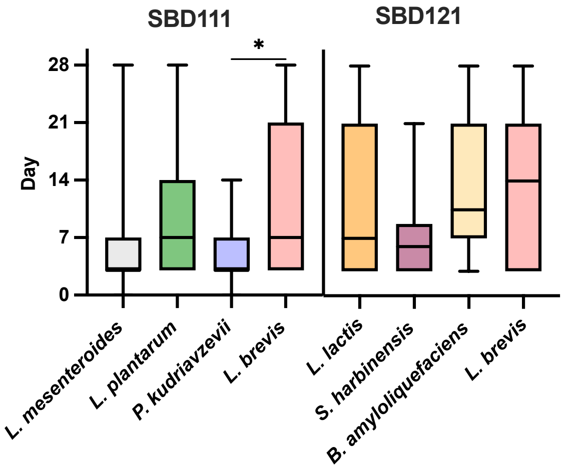


**Figure S1.** Time to non-detection of probiotic strains. Box and whisker plots showing the time (in days) until each of the administered probiotic strains was no longer detectable in stool samples via qPCR. Data were analyzed using one-way ANOVA with Tukey's multiple comparison test; *$p < 0.05$.

**Figure S2.** Analysis of microbial diversity.

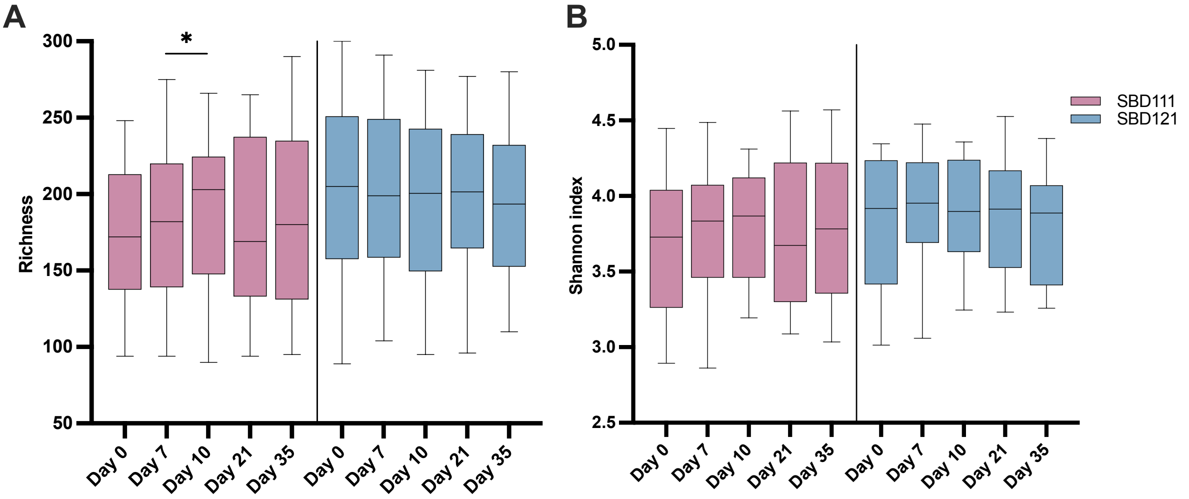


**Figure S2.** Analysis of microbial diversity. Box plots comparing **A.** bacterial species richness and **B.** Shannon diversity indices between Day 7 and other time points (baseline, Day 10, 21, and 35) for each study group. Box plots show the median, interquartile range (IQR), and 95% confidence intervals (CI). Statistical significance was assessed using a linear mixed-effects regression model. Asterisks denote significance levels: *p < 0.05.

**Figure S3.** Analysis of functional diversity.

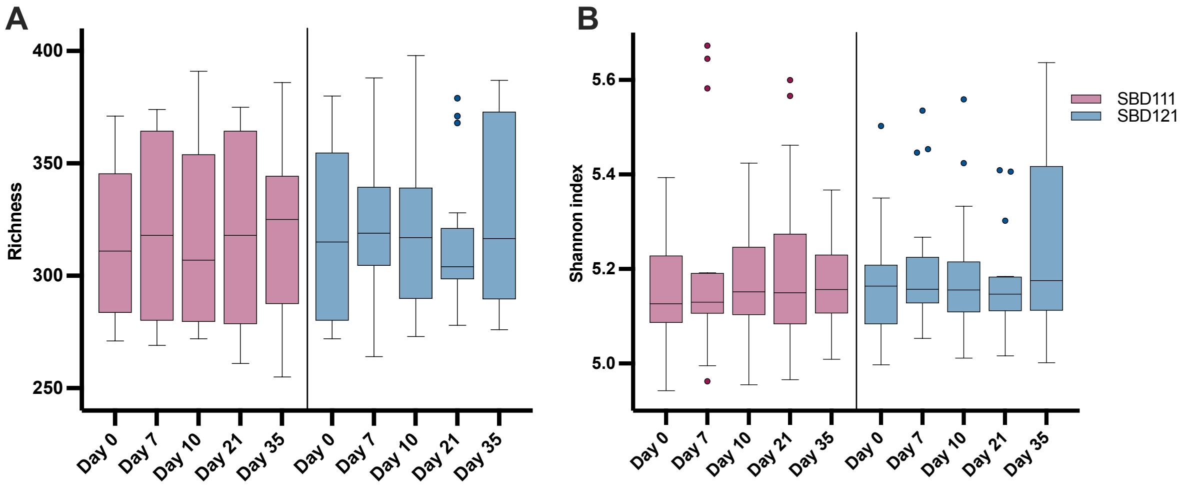


**Figure S3.** Analysis of functional diversity. Box plots comparing **A.** MetaCyc functional pathway richness and **B.** MetaCyc functional pathway Shannon diversity indices between day 7 and other time points (baseline, Day 10, 21, and 35) for each study group. Box plots show the median, interquartile range (IQR), and 95% confidence intervals. Statistical significance was assessed using a linear mixed-effects regression model.

**Figure S4.** Box plots comparing the relative abundance of synbiotic medical food strains.

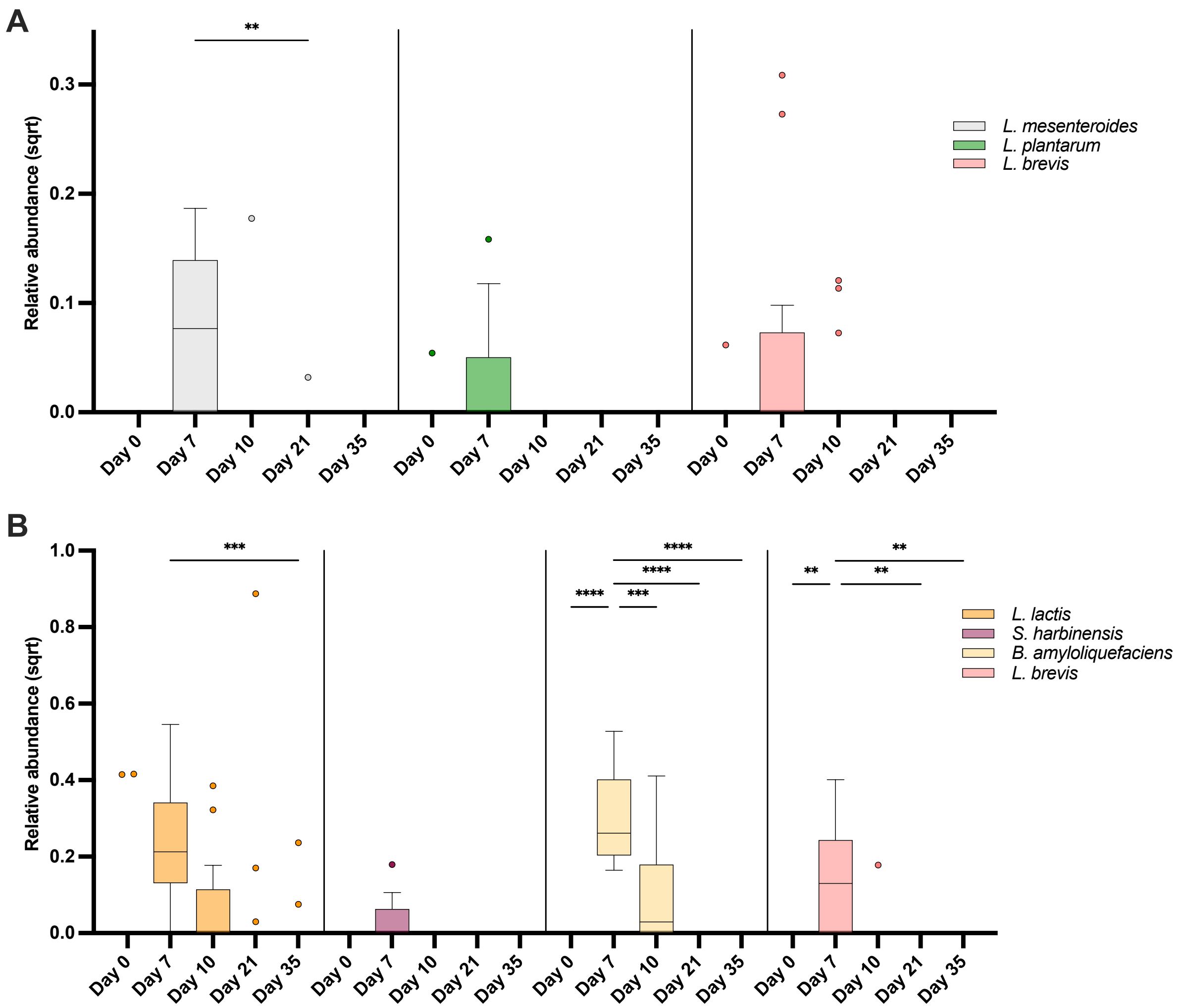


**Figure S4.** Box plots comparing the relative abundance of synbiotic medical food strains. **A.** SBD111 strains (*L. mesenteroides*, *L. plantarum*, and *L. brevis*) and **B.** SBD121 strains (*L. brevis*, *B. amyloliquefaciens*, *S. harbinensis*, and *L. lactis*) in stool metagenomes between Day 7 and other time points (baseline, Day 10, 21, and 35). Relative abundance values were estimated using MetaPhlAn marker genes. *P. kudriavzevii* could not be reliably detected in the metagenomes, likely due to the paucity of clade-specific marker genes available for this species in the MetaPhlAn database, particularly at low abundance values. Box plots depict median, IQR, and 95% CI. P-values, **p<0.01, ***p<0.001, and ****p<0.0001, represent significance estimated from linear mixed effects regression model interaction terms after FDR correction.

**Figure S5.** Median colony forming units (CFU) per gram of feces across different media types.

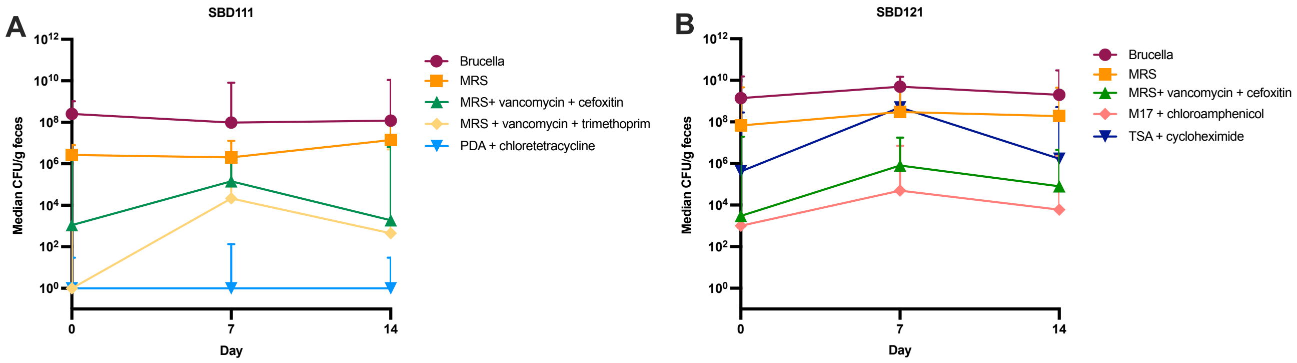


**Figure S5.** Median colony forming units (CFU) per gram of feces across different media types. Line plot showing microbial counts recovered from fecal samples using culture-based plating on various selective and non-selective media. Data are shown as median CFU/g across participants at baseline, after seven days of product consumption, and one-week post-consumption time points. One-way ANOVA with Tukey's multiple comparisons test was conducted, but no statistically significant differences were observed.
